# Neuronal avalanches in temporal lobe epilepsy as a diagnostic tool: a noninvasive investigation of intrinsic resting state dynamics

**DOI:** 10.1101/2023.12.06.23299589

**Authors:** Marie-Constance Corsi, Emahnuel Troisi Lopez, Pierpaolo Sorrentino, Alberto Danieli, Simone Cuozzo, Palo Bonanni, Gian Marco Duma

**Affiliations:** Sorbonne Université, Institut du Cerveau – Paris Brain Institute -ICM, CNRS, Inria, Inserm, AP-HP, Hopital de la Pitié Salpêtrière, F-75013, Paris, France; Institute of Applied Sciences and Intelligent Systems of National Research Council, Pozzuoli, Italy; Institut de Neurosciences des Systèmes, Aix-Marseille Université, 13005 Marseille, France; University of Sassari, Department of Biomedical Sciences, Viale San Pietro, 07100, Sassari, Italy; IRCCS E. Medea Scientific Institute, Epilepsy Unit, Via Costa Alta 37, 31015, Conegliano, Treviso, Italy

**Author notes:** Corresponding author: Pierpaolo Sorrentino, Institut de Neurosciences des Systèmes, Aix-Marseille Université, 13005 Marseille, France. Co-First authors (Marie-Constance Corsi, Emahnuel Troisi Lopez). Authors email: Marie-Constance Corsi, Emahnuel Troisi Lopez, Gian Marco Duma.

## Abstract

**Background and Objectives:** The epilepsy diagnosis still represents a complex process, with misdiagnosis reaching 40%. Here, we aimed at building an automatable workflow, to help the clinicians in the diagnostic process, differentiating between controls and a population of patients with temporal lobe epilepsy (TLE). While primarily interested in correctly classifying the participants, we used data features providing hints on the underlying pathophysiological processes. Specifically, we hypothesized that neuronal avalanches (NA) may represent a feature that encapsulates the rich brain dynamics better than the classically used functional connectivity measures (Imaginary Coherence; ImCoh).

**Methods:** We recorded 10 minutes of resting state activity with high-density scalp electroencephalography (hdEEG; 128 channels). We analyzed large-scale activation bursts (NA) from source activation, to capture altered dynamics. Then, we used machine-learning algorithms to classify epilepsy patients vs. controls, and we described the goodness of the classification as well as the effect of the durations of the data segments on the performance.

**Results:** Using a support vector machine (SVM), we reached a classification accuracy of 0.87 ± 0.10 (SD) and an area under the curve (AUC) of 0.94 ± 0.06. The use of avalanches-derived features, generated a mean increase of 16% in the accuracy of diagnosis prediction, compared to ImCoh. Investigating the main features informing the model, we observed that the dynamics of the entorhinal cortex, superior and inferior temporal gyri, cingulate cortex and prefrontal dorsolateral cortex were informing the model with NA. Finally, we studied the time-dependent accuracy in the classification. While the classification performance grows with the duration of the data length, there are specific lengths, at 30s and 180s at which the classification performance becomes steady, with intermediate lengths showing greater variability. Classification accuracy reached a plateau at 5 minutes of recording.

**Discussion:** We showed that NA represents a better EEG feature for an automated epilepsy identification, being related with neuronal dynamics of pathology-relevant brain areas. Furthermore, the presence of specific durations and the performance plateau might be interpreted as the manifestation of the specific intrinsic neuronal timescales altered in epilepsy. The study represents a potentially automatable and noninvasive workflow aiding the clinicians in the diagnosis.

## 1. Introduction

Epilepsy is one of the most diffused neurological conditions, affecting more than 50 million people worldwide, representing a relevant burden to health services (Juarez-Garcia et al., 2006). Even if consensus statements and international guidelines are present, the diagnosis of epilepsy still represents a complex scenario, with a rate of misdiagnosis, which varies widely across different centers, reaching 40% in specific cases (Uldall et al., 2006; Zaidi et al., 2000). In this context, medical doctors may need support in the diagnostic process (Oto, 2017). Recent years have witnessed the introduction of artificial intelligence (AI) methodologies as a supporting tool for clinicians both in the diagnostic and treatment processes. In the field of epilepsy, vast efforts were conducted in order to translate the large amount of multimodal data that are nowadays collected during the clinical practice, towards the improvement, and potentially the automation, of the diagnostic process (Ilias et al., 2023; Jin et al., 2021; Spitzer et al., 2022). Across the multiple neuroimaging modalities, electroencephalography (EEG) represents the election tool in epilepsy diagnosis (Tatum et al., 2018). The vast majority of applications of AI on the EEG signal have focused on the automatic recognition of epileptiform/seizure activity or seizure forecasting (Acharya et al., 2018; Tveit et al., 2023; Zhang et al., 2020; Zhou et al., 2018). However, the simple but fundamental diagnostic question, namely the identification of epileptic patients, is far from being efficiently addressed in an automated fashion. Very few studies exploited AI trying to provide an automatic tool aiding the clinician in the epilepsy diagnosis (Rijnders et al., 2022; Varone et al., 2021; Wang et al., 2022). In fact, these investigations displayed a large variability in the accuracy and sensitivity which can be related to the EEG features used to train the model, opening the question of which is the feature of the EEG signal better suited for training an AI model for diagnosis. Until now, most of the studies attempting to perform AI-based diagnosis exploited functional connectivity in the sensor space, derived by the covariation between signals and/or spectral power from electrodes activity, as well as the frequency-specific phase locking (i.e. synchronization) (Saeidi et al., 2021). However, such techniques rely on the assumption of stationarity of the EEG signals, which may represent an important limitation, and might affect the stability of the estimates. In the present work, we aimed at identifying a better suited EEG signal feature to train an AI-model-based classification (epilepsy vs. non-epilepsy). We hypothesized that the aperiodic activation bursts (neuronal avalanches) of source-derived EEG activities may represent a better measure for an AI-model, since they are not based on the assumptions of stationarity. In particular, neuronal avalanches (NA) belong to the framework of criticality, which lends itself nicely to the investigation of the microscopic dynamical behavior of neuronal assemblies and its (non-linear) relation to large-scale alterations such as, in the case of epilepsy, seizure initiation, propagation and termination (Hagemann et al., 2021; Liu et al., 2023; Moosavi & Truccolo, 2023). Recent findings suggest that NA spreading on the large scale represents a more stable and reliable feature for the individualized investigation and task classification of brain dynamics (Corsi et al., 2023; Polverino et al., 2022; Sorrentino, Troisi Lopez, et al., 2023). Relevantly, our previous work has highlighted the altered spreading of NA and its relation to brain morphology in temporal lobe epilepsy (TLE), using the source-derived high-density EEG (hdEEG) activity at rest (Duma et al., 2023). In the light of this, here we hypothesized that the spreading of the aperiodic activation bursts at the whole-brain level may represent a relevant feature towards the automatic classification of epileptic vs. non-epileptic patients. To this end, we recorded resting-state high-density EEG (hdEEG) from patients with temporal lobe epilepsy and a control group. The model of TLE is of particular interest as it represents a rather well-defined group of electroclinical conditions with lower clinical heterogeneity compared to other epilepsy forms. Using source-reconstructed EEG signals, we individuated the presence of NA and used them to calculate the probability of consecutive activations between brain regions (i.e., the topographical spreading of the avalanches). This information was stored within an adjacency matrix named avalanche transition matrix (ATM) (Sorrentino et al., 2022) which was then used as a feature for a support vector machine (SVM) classifier. We hypothesized that ATMs would have performed better, compared to the power-based connectivity metric generally used in literature, differentiating with a larger accuracy the two conditions (epilepsy vs. non-epilepsy). Relevantly, we computed the ATM from a resting state activity free from epileptiform activity, in order to investigate if the basal functional organization of the system, without the presence of characteristic graphic-elements (i.e., seizure and/or interictal epileptiform discharges), may contribute to a reliable classification of the patients. We then looked into the most relevant feature pattern, namely the most informative elements used by the model to differentiate the two groups. We expected that the dynamics of those areas linked with the pathophysiology of TLE, would have informed the model more than other brain regions. This endows our results with interpretability and allows the identification of clusters of regional interactions that might be relevant to the pathology. Finally, we investigated the classification performance with respect to the duration of the brain signals. Hence, we have performed the classification based on the growing amount of data used to compute ATMs, ranging from 5 to 300 sec. To summarize, the purpose of the present work is to exploit the spreading of the aperiodic activities at the whole-brain level as a feature to inform an AI-based model aimed at the automatic identification of epileptic patients, in order to provide the clinicians with a practical and non-invasive tool supporting the diagnostic process.

## 2. METHOD

### 2.1 Participants

We retrospectively enrolled 70 patients with temporal lobe epilepsy, who underwent high-density electroencephalography (hdEEG) for clinical evaluation between 2018-2021 at the Epilepsy and Clinical Neurophysiology Unit, IRCCS Eugenio Medea cited in Conegliano (Italy). The diagnostic workflow included clinical history and examination, neuropsychological assessment, long-term surface Video EEG (32 channels) monitoring, hdEEG resting-state recording, brain magnetic resonance imaging and positron emission tomography (PET) as an adjunctive investigation in selected cases. The diagnosis of temporal lobe epilepsy was established according to the ILAE guidelines. We selected for this specific work a subgroup of 31 patients with left temporal lobe epilepsy (mean age = 37.18 [SD = 18.04]; 14Lfemales). The control group was composed of 31 healthy participants with no history of neurological or psychiatric disorders (mean age = 34.92 [SD = 9.22]; 25Lfemales). A description of patients’ demographic and clinical characteristics is provided in Table 1. The study protocol was conducted according to the Declaration of Helsinki and approved by the local ethical committee.

**TABLE 1.**
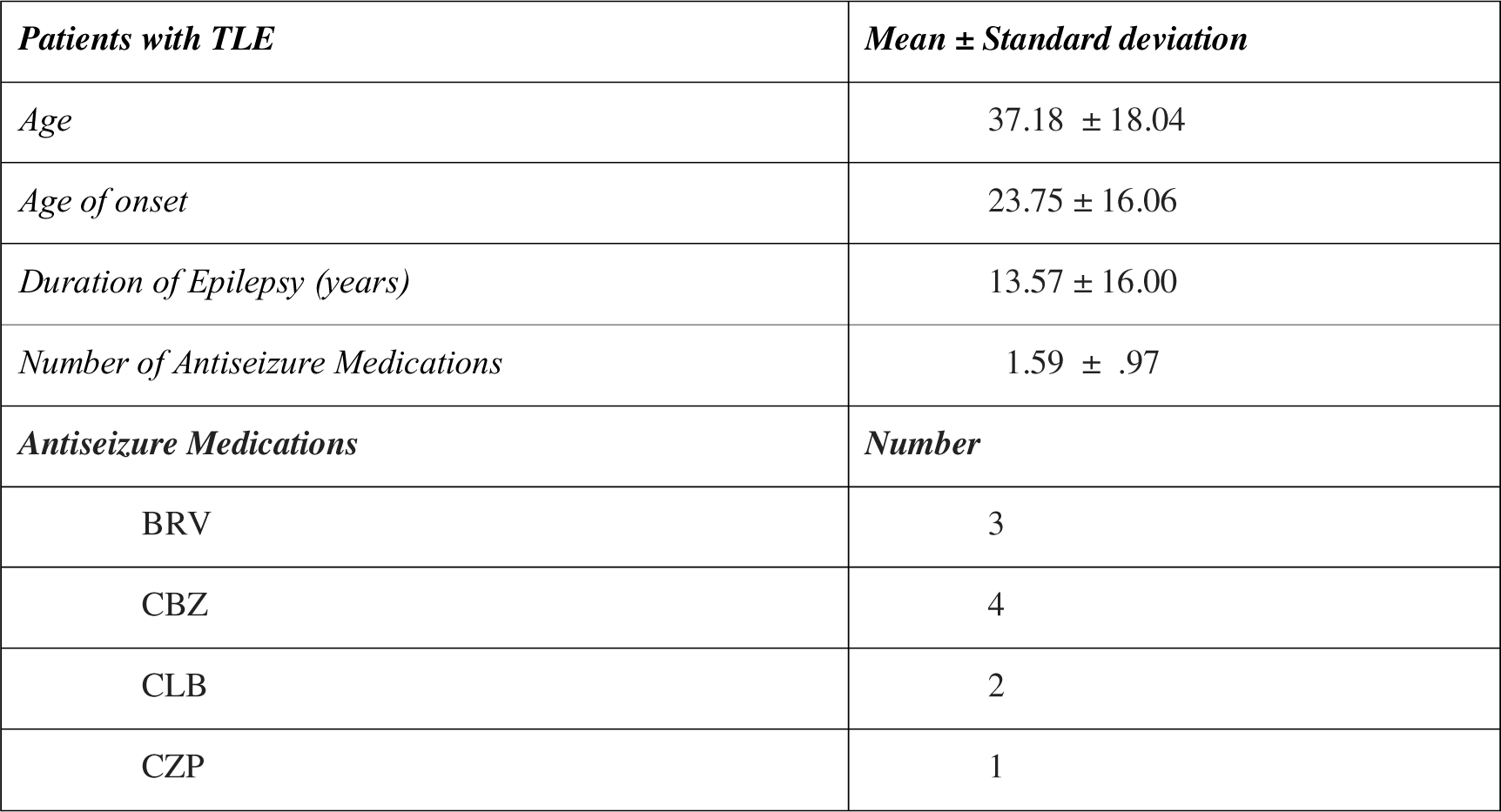
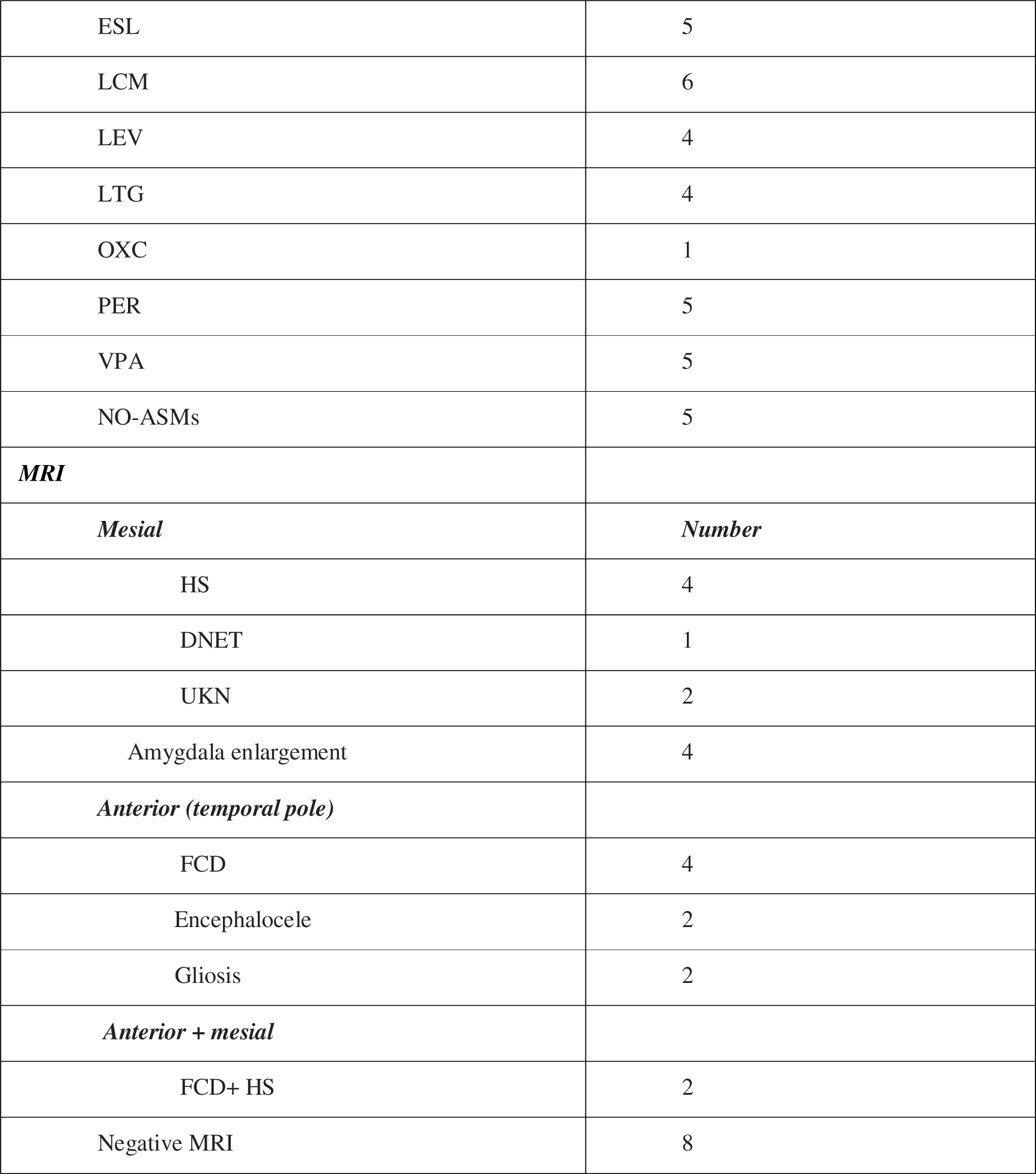
The table describes the demographic and clinical characteristics of the patients with temporal lobe epilepsy. MRI abnormalities are reported by sublobar localization. The continuous variables are reported as mean ± standard deviation. Antiseizure Medication abbreviations: BRV = brivaracetam, CBZ = carbamazepine, CLB = clobazam, CZP = clonazepam, ESL= eslicarbazepine, LCM = lacosamide, LEV = levetiracetam, LTG = lamotrigine, OXC = oxcarbazepine, PB = phenobarbital, PER = perampanel, VPA= valproic acid, NO-ASMs = no pharmacological treatment. Abbreviation of the identified anomalies on the magnetic resonance imaging: FCD= focal cortical dysplasia, HS=hippocampal sclerosis, DNET = dysembryoplastic neuroepithelial tumors, UKN = unknown.

**TABLE 2.**
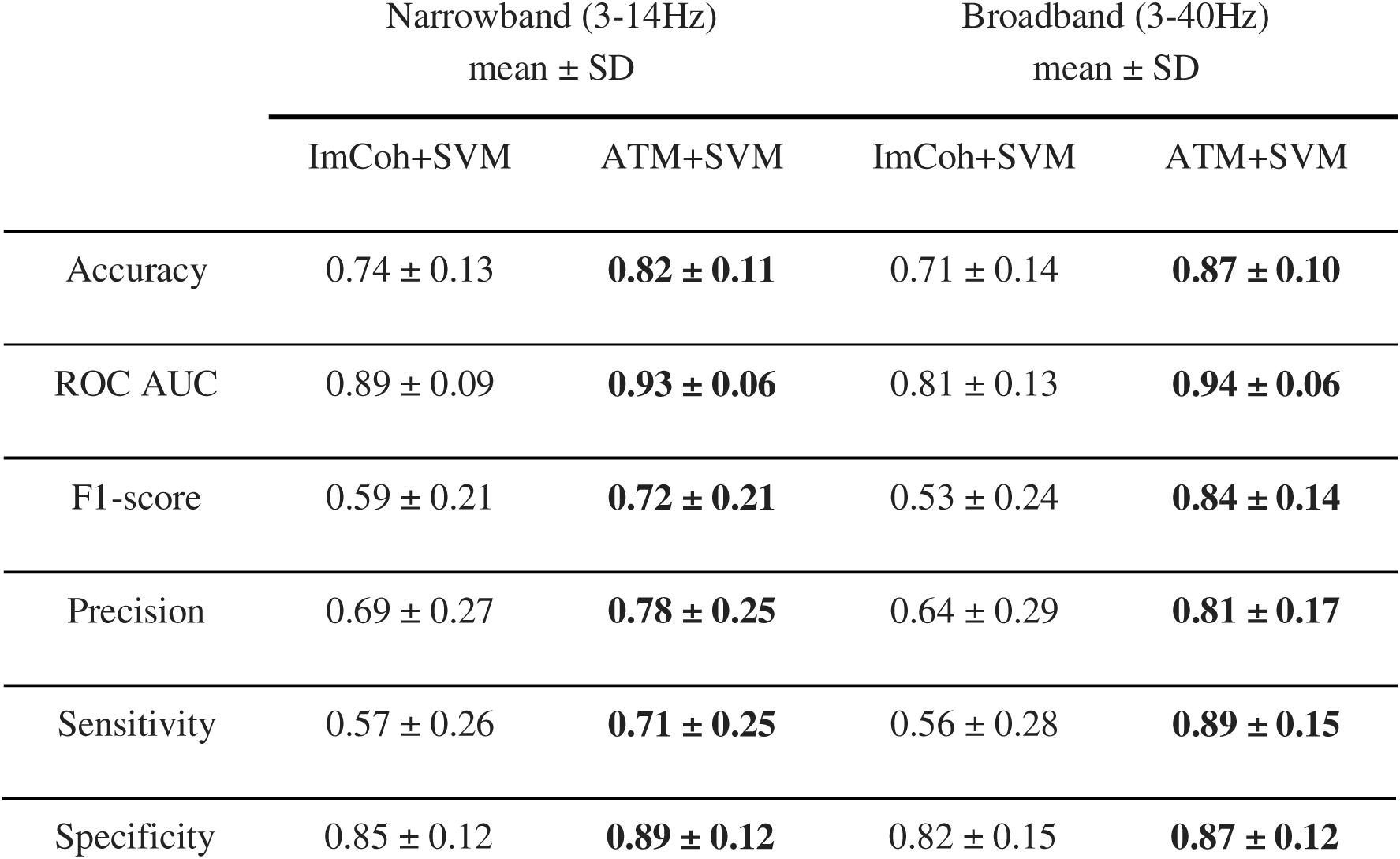
Classification performance. . The present table displays the indices of the classification performance when considering the model trained with avalanche transition matrix (ATM) or imaginary coherence (ImCoh) obtained from the narrowband (3-14Hz) or broadband (3-40Hz) signal.

| | Narrowband (3-14Hz)<br>mean ± SD | | Broadband (3-40Hz)<br>mean ± SD | |
| --- | --- | --- | --- | --- |
|  | ImCoh+SVM | ATM+SVM | ImCoh+SVM | ATM+SVM |
| Accuracy | $0.74 \pm 0.13$ | **$0.82 \pm 0.11$** | $0.71 \pm 0.14$ | **$0.87 \pm 0.10$** |
| ROC AUC | $0.89 \pm 0.09$ | **$0.93 \pm 0.06$** | $0.81 \pm 0.13$ | **$0.94 \pm 0.06$** |
| F1-score | $0.59 \pm 0.21$ | **$0.72 \pm 0.21$** | $0.53 \pm 0.24$ | **$0.84 \pm 0.14$** |
| Precision | $0.69 \pm 0.27$ | **$0.78 \pm 0.25$** | $0.64 \pm 0.29$ | **$0.81 \pm 0.17$** |
| Sensitivity | $0.57 \pm 0.26$ | **$0.71 \pm 0.25$** | $0.56 \pm 0.28$ | **$0.89 \pm 0.15$** |
| Specificity | $0.85 \pm 0.12$ | **$0.89 \pm 0.12$** | $0.82 \pm 0.15$ | **$0.87 \pm 0.12$** |

### 2.2 Resting State EEG recording

The hdEEG recordings were obtained using a 128-channel Micromed system referenced to the vertex. Data was sampled at 1,024 Hz and the impedance was kept below 5kΩ for each sensor. For each participant, we recorded 10 minutes of closed-eyes resting state while comfortably sitting on a chair in a silent room.

### 2.3 EEG pre-processing

Signal preprocessing was performed via EEGLAB 14.1.2b (Delorme & Makeig, 2004). The continuous EEG signal was first downsampled at 250 Hz and then bandpass-filtered (0.1 to 45 Hz) using a Hamming windowed sinc finite impulse response filter (filter order = 8250). The signal was visually inspected to identify interictal epileptiform discharges (IEDs) by the clinicians and then segmented into 1-sec-long epochs. Epochs containing IEDs activities were removed. We purposely removed epochs containing IEDs since we wanted to use the intrinsic brain functional organization of the brain, independent from epileptiform activity, for patients vs. control classification. The epoched data underwent an automated bad-channel and artifact detection algorithm using the TBT plugin implemented in EEGLAB. This algorithm identified the channels that exceeded a differential average amplitude of 250μV and marked those channels for rejection. Channels that were marked as bad in more than 30% of all epochs were excluded. Additionally, epochs having more than 10 bad channels were excluded. We automatically detected possible flat channels with the Trimoutlier EEGLAB plug-in within the lower bound of 1μV. We rejected an average of 40.15 ± 41.24 (SD) epochs. The preprocessing analysis pipeline has been applied by our group in previous studies investigating both task-related and resting state EEG activity (Duma et al., 2021, 2022). Data cleaning was performed with independent component analysis (Stone, 2002), using the Infomax algorithm (Bell & Sejnowski, 1995) as implemented in EEGLAB. The resulting 40 independent components were visually inspected and those related to eye blinks, eye movements, muscle, and cardiac artifacts were discarded. An average of 9.51± 4.68 (SD) components were removed. The remaining components were then projected back to the electrode space. Finally, bad channels were reconstructed with the spherical spline interpolation method (Perrin et al., 1989). The data were then re-referenced to the average of all electrodes. At the end of the data preprocessing, each subject had at least 6 minutes of artifact-free signal.

### 2.4 Cortical Source modeling

We used the individual anatomy MRI to generate individualized head models for the patients with TLE. The anatomic MRI for source imaging consisted of a T1 isotropic three-dimensional (3D) acquisition. For twelve patients and for the control group we used the MNI-ICBM152 default anatomy (Evans et al., 2012) from Brainstorm (Tadel et al., 2011) since the 3D T1 MRI sequences were not available. The MRI was segmented into skin, skull, and gray matter using the Computational Anatomy Toolbox (CAT12) (Dahnke et al., 2013). The resulting individual surfaces were then imported in Brainstorm, where three individual surfaces adapted for Boundary Element Models (BEM) were reconstructed (inner skull, outer skull and head) and the cortical mesh was downsampled at 15,002 vertices. The co-registration of the EEG electrodes was performed using Brainstorm by projecting the EEG sensor positions on the head surface with respect to the fiducial points of the individual or the template MRI. We applied manual correction of the EEG cap on the individual anatomy whenever needed, prior to projecting the electrodes on the individual head surface. We then derived an EEG forward model using the 3-shell BEM model (conductivity: 0.33, 0.165, 0.33 S/m; ratio: 1/20) (Goncalves et al., 2003) estimated using OpenMEEG method implemented in Brainstorm (Gramfort et al., 2010). Finally, we used the weighted minimum norm imaging (Hämäläinen & Ilmoniemi, 1994) as the inverse model, with the Brainstorm’s default parameters setting.

### 2.5 Features extraction

#### 2.5.1. Avalanche transition matrix computation

Similarly to our previous work using neuronal avalanches (Duma et al., 2023), we extracted the activity of a total of 68 regions of interest (ROIs) from the Desikan-Killiany atlas (Desikan et al., 2006). The ROIs time series were obtained by averaging the activity across the vertices composing each ROI. To study the dynamics of brain activity, we estimated “neuronal avalanches” from the source-reconstructed ROI time series. Firstly, the time series of each ROI was discretized by calculating the z-score over time and then setting positive and negative excursions beyond a threshold as 1, and the rest of the signal as 0 (Sorrentino et al., 2021). A neuronal avalanche begins when, in a sequence of contiguous time bins, at least one ROI is active (i.e., above threshold), and ends when all ROIs are inactive (Beggs & Plenz, 2003; Shriki et al., 2013; Sorrentino et al., 2021). These analyses require the time series to be binned. This is done to ensure that one is capturing critical dynamics, if present. To estimate the suitable time bin length, for each subject, each neuronal avalanche, and each time bin duration, the branching parameter σ was estimated (Haldeman & Beggs, 2005). In fact, systems operating at criticality typically display a branching ratio ∼1. The branching ratio is calculated as the geometrically averaged (over all the time bins) ratio of the number of events (activations) between the subsequent time bin (descendants) and that in the current time bin (ancestors), and then averaging over all the avalanches (Bak et al., 1987). More specifically:

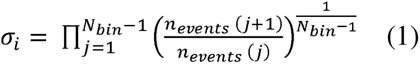

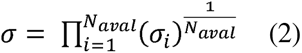

where σ *_i_* is the branching parameter of the *ith* avalanche in the dataset, *N_bin_*is the total amount of bins in the *i-th* avalanche, *n_events_ (j)* is the total number of events active in the *j-th* bin, and *N_aval_*is the total number of avalanche in the dataset. In our analyses, the branching ratio was 1 for bin = 1 (corresponding to bins of 4 ms). An avalanche-specific transition matrix (ATM) was calculated where element (*i, j*) represented the probability that region j was active at time *t +*δ, given that region *i* was active at time ***t***, where δ ∼ 4 ms. The ATMs were averaged element-wise across all avalanches per each participant. It is important to mention that the ATMs were computed over the broadband signal. However, given the relevance of the slow frequencies (i.e., theta and alpha bands) in the TLE (Scherer et al., 2020; Sip et al., 2021), we also computed ATMs on a narrowband filtered signal (theta and alpha bands; 3-14 Hz). This was performed to evaluate the potential increase of the accuracy of classification when including only specific frequency bands. For each tested frequency band, to ensure that we were in the best condition to reach the highest classification performance possible, we optimized the threshold applied to the z-scored signals (ranging from 1.2 to 3.0). Finally, we computed the ATMs multiple times, each time averaging over different lengths of the signals (i.e., 5, 15, 30, 60, 120, 180, and 300sec). This was performed in order to investigate the effect of signal length used in the ATMs computation on the classification performance. To mitigate the influence of the position of a given temporal window aJ in the registration on the classification performance, we randomly picked 100 times aJ across the registration, and we considered the median value of the classification accuracy over the permutations.

#### 2.5.2. Imaginary Coherence

Additionally to the ATMs, we computed the imaginary coherence in order to compare the performance with a widely used metric of functional connectivity. The imaginary part of the coherence (ImCoh) has been demonstrated to be robust to volume conduction, improving the identification of true interactions in EEG-derived functional connectivity (Haufe et al., 2013; Haufe & Ewald, 2019; Nolte et al., 2004). We computed ImCoh as it follows:

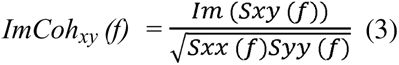

where *S_xy_* is the cross-spectral density between region *x* and region *y*, while *S*_xx_ and *S*_yy_ represent the autospectral density of the two regions. The spectral densities were estimated using a multitaper method with digital prolate spheroidal sequence (DPSS) (Moore & Cada, 2004) windows of 10s, an overlap of 50%, and a maximum frequency resolution of 0.1Hz. To be consistent with the scheme used with the ATMs, we calculated the ImCoh over both the broadband and the narrowband filtered signal (3-14 Hz).

### 2.6. Classification pipeline

To perform the classification, we used a Support Vector Machine (SVM) classifier (Smola & Schölkopf, 2004). We optimized the SVM with a 5-fold cross-validated grid search to find optimal parameters, namely the kernels (linear or Radial Basis Function-RBF) and the regularization hyperparameter C during the training step. The kernel functions enable the system to work in a high-dimensional space and to allow for more complex decision functions when the data is not linearly separable (Faouzi & Colliot, 2023). The C parameter adjusts the misclassification of training examples against the simplicity of the decision. The classification scores were evaluated by cross-validation consisting of a random permutation cross-validator with a training size of 80%, a test size of 20%, and 50 re-shuffling and splitting iterations. To assess the performance of the classification we reported several metrics, namely the accuracy classification score, the Area Under the Receiver Operating Characteristic Curve (ROC AUC) from the prediction scores, the F1, the precision, the sensitivity, and the specificity scores. To investigate the interpretability of the classification performance, we studied the relative importance of the features, derived from the absolute value of the classification coefficients of the classification model. Here, we computed the median value over the splits of the cross-validation of the testing set. The Python packages scikit-learn (Buitinck et al., 2013), MNE-python (Gramfort, 2013), SciPy (Virtanen et al., 2020), NumPy (Harris et al., 2020) and Pandas (McKinney, 2010) were used to perform the analysis and to compute the results. The code is publicly available at: https://github.com/mccorsi/NeuronalAvalanches_TemporalLobeEpilepsy_EEG. We performed classification training on the model with ATMs and ImCoh features.

## 3. Results

### 3.1 Model prediction performance

The implemented SVM model classified the participants as TLE or controls based on the global system dynamics, namely the ATMs, computed on the broadband signal (3-40Hz), with a mean accuracy of (0.87 ± 0.10). We observed that the prediction accuracy of the model trained on the ATMs was larger as compared to the one trained with ImCoh (0.71 ± 0.14). Importantly, these results were also confirmed for the signals filtered in the narrowband (3-14 Hz) (ATMs accuracy: 0.82 ± 0.11; ImCoh accuracy: 0.74 ± 0.13). The better classification performance for ATMs vs. ImCoh was also confirmed by an increased area under the curve (AUC), both when considering the broadband and the narrowband filtered signal. In general, the results display a better classification across all the performance indices in the model trained with the ATMs as compared to the ImCoh (see Table.1).

We note that, as suggested by recent findings, providing exclusively the mean across splits of the classification accuracies might be misleading for the interpretation of the model performance (Shafiezadeh et al., 2023). Hence, here we display the full distribution of the classification accuracies across the multiple splits of the cross-validation (see Fig.2A). Notably, the distribution of the prediction accuracies with the ATMs is narrower, and skewed towards larger accuracy values. Additionally, in Fig.2, we also provide the mean receiver operating characteristic (ROC) curve and the corresponding AUC, providing further evidence of the increased performance of the ATMs compared to the ImCoh. In fact, in the supplementary material, we also provide the ROC curves across cross-validation splits (see Supplementary Figure 1). However, in the manuscript, we display for simplicity the mean ROC across splits. To summarize, both broadband and narrow-band filtered signals display similar behavior, namely a better performance when using the aperiodic part of the signal as opposed to using a metric that assumes stationarity and is purely power-based. From here on we will limit ourselves to reporting the results in the broadband signal. However, the results in theta-alpha band signal are shown in the supplementary materials.

**FIGURE 1.**
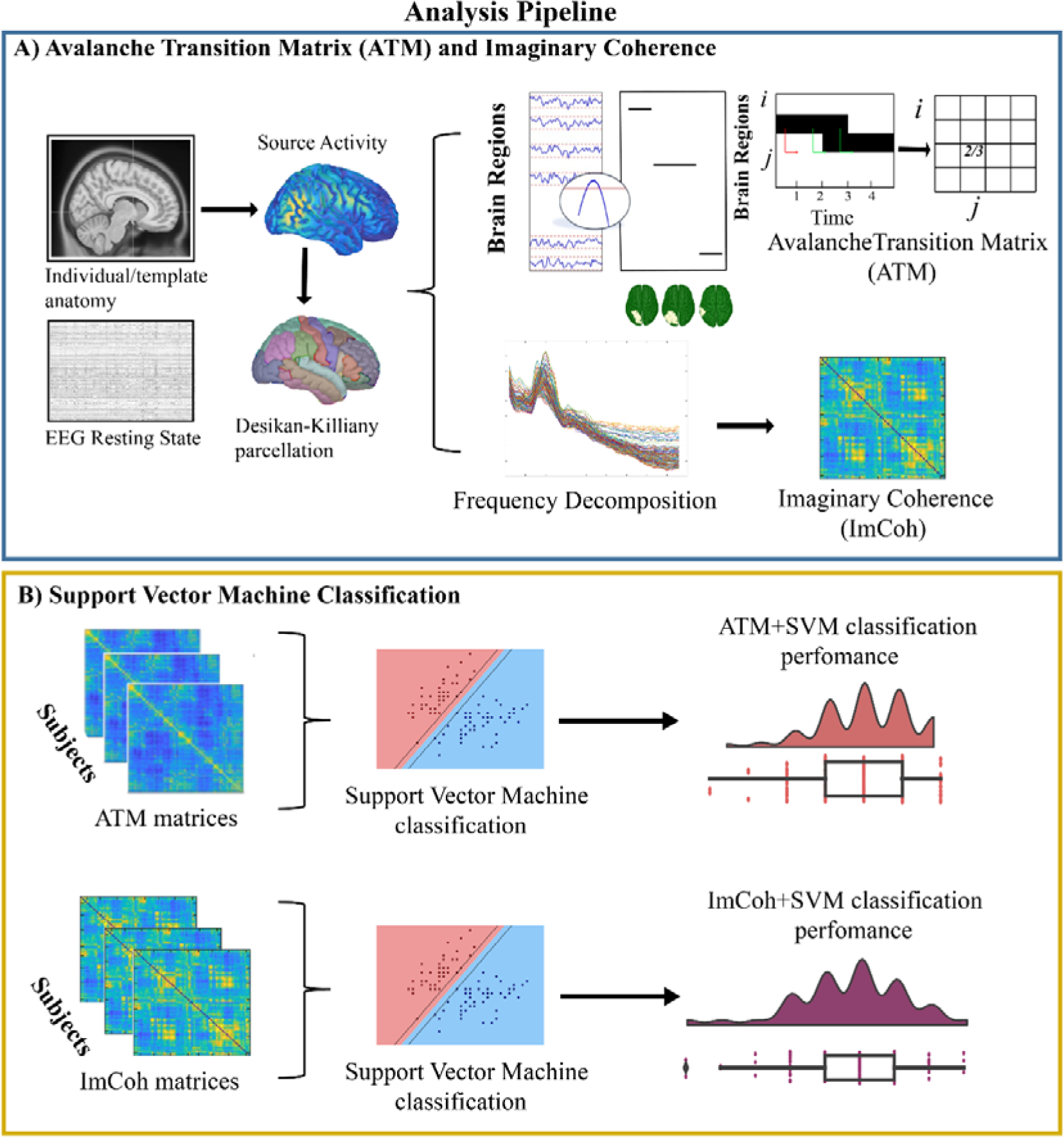
Analysis pipeline. The present figure illustrates the analytical steps used for feature extraction and machine learning based classification via support vector machine.

**FIGURE 2.**
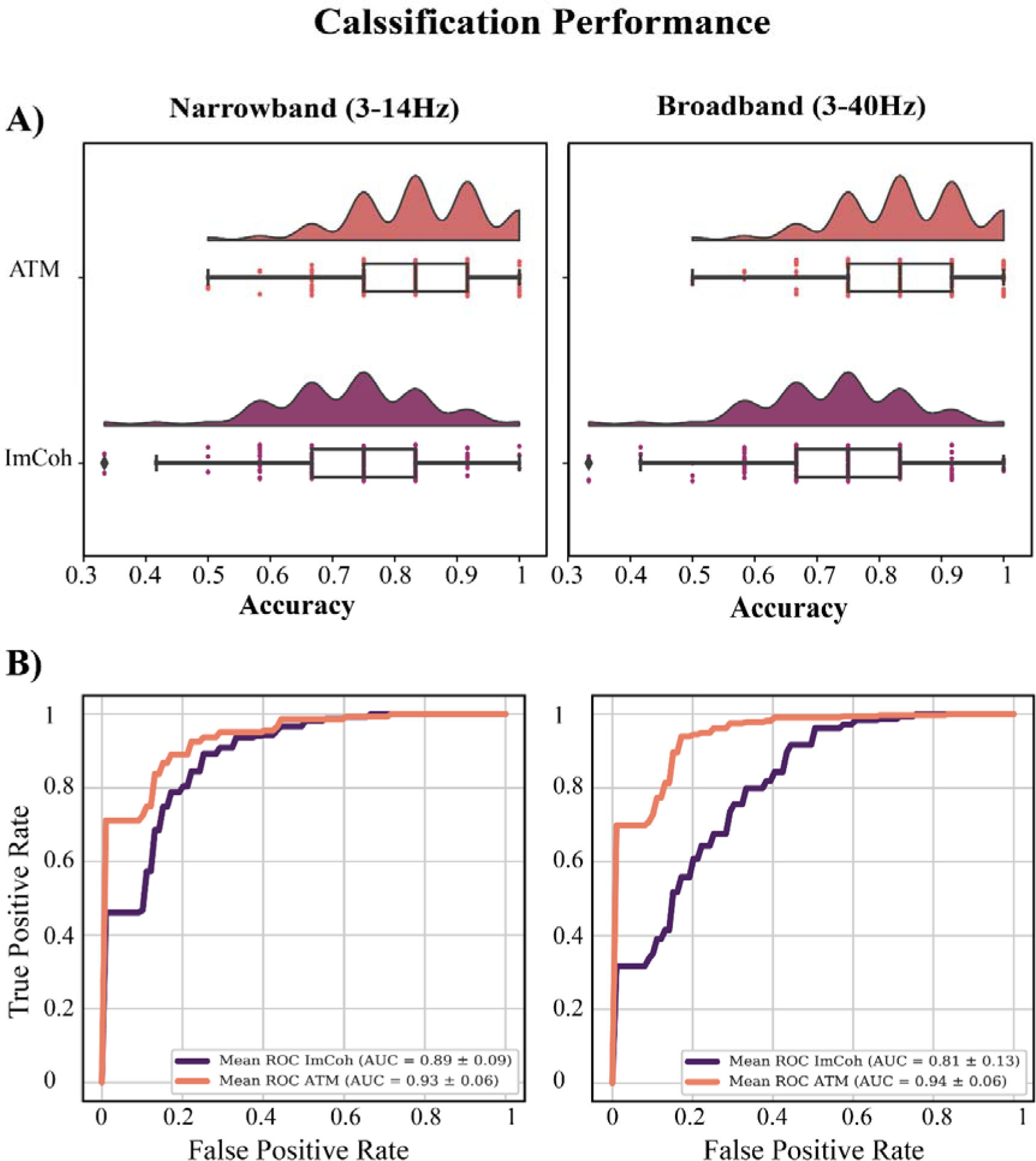
Accuracy and receiver operating characteristic curves. Panel A of the present picture displays the full distribution of the classification accuracy of the support vector machine (SVM) model across the cross-validation splits, when trained with avalanche transition matrix (ATM, in salmon) and imaginary coherence (ImCoh, in purple). On the left the accuracy classification using narrow-band filtered signal (3-14 Hz) and on the right the broadband signal (3-40 Hz). Panel B shows the mean receiver operating characteristic (ROC) curves across cross-validation splits and the corresponding area under the curve (AUC), both for ATM (salmon line) and ImCoh (purple line).

### 3.2 Feature selected by the model

A relevant topic in automatic learning is related to interpretability, that is the ability to trace back the specific information that is being used to classify items. In our case, our input data was represented by a node-by-node matrix, where each edge indicates the power of the cross-covariance (ImCoh) or the transition probability (ATMs) between two nodes. We observed a difference in the features used by the model to classify between TLE patients and healthy controls between the functional measures we used. When considering the ImCoh, we observed that the distribution of the relevant features is narrow, which means that most of the edges hold similar importance for the classification. Conversely, we observed a larger distribution of the importance of the features in the ATMs-based classification, which demonstrates that a specific subset of edges carries the information that is used to differentiate the two groups (see Fig.3A). This difference is also visualized in the chord diagrams in Fig.3B. One can observe similar importance values across most of the connections when considering ImCoh while, in the case of the ATMs, specific edges emerge from the background. The edge’s relevance appeared to be clustered into specific nodes, which were different between ATM and ImCoh. On the one hand, when considering the model trained with the ATMs, the features with the largest importance were clustered around brain regions of fundamental relevance in TLE, involved both in seizure initiation and/or propagation, or characterized by well-known metabolic or connectivity alterations, such as the entorhinal cortex, the superior and inferior temporal gyri, the cingulate cortex and the prefrontal dorsolateral cortex (Bernasconi et al., 1999; Jo et al., 2019; Kubota et al., 2013; Qin et al., 2020; Vismer et al., 2015). On the other hand, the model with ImCoh highlighted as more important the edge’s clustering on pre- and postcentral gyri and rostral middle frontal regions.

**FIGURE 3.**
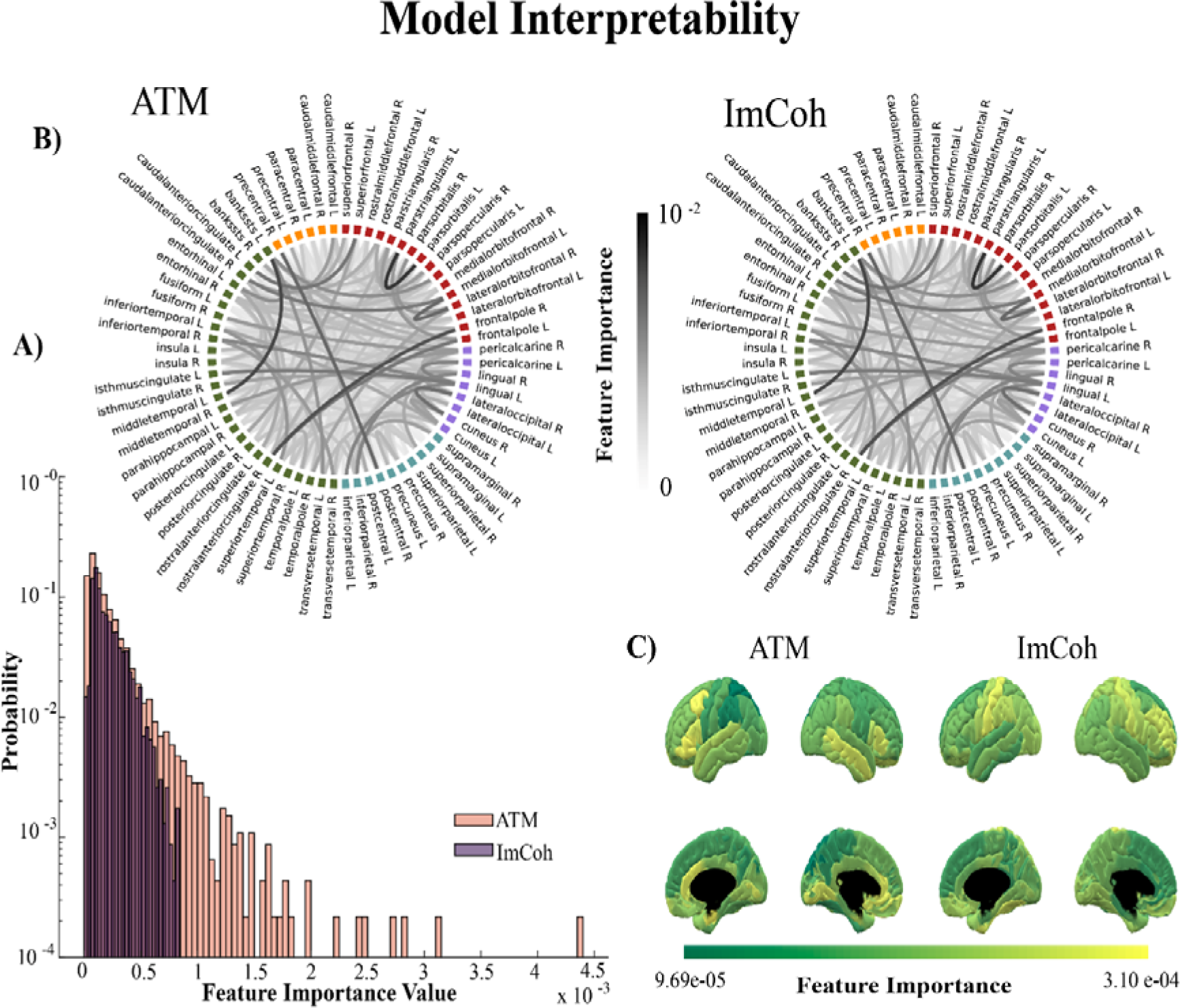
Feature importance for model interpretability. The present figure represents the relevance of the information used by the model to perform classification, namely the feature importance, both using avalanche transition matrix (ATM) and imaginary coherence (ImCoh). Panel A is a histogram of the probability of the feature importance value in ATM and ImCoh. The histogram shows a narrow distribution for ImCoh and a broader distribution for ATM, suggesting that in ATMcertain edges drive the majority of information necessary for differentiating the two groups. Panel B is the edge representation in a chord plot, showing the importance of each edge in the classification. Finally, panel C is the mean importance value of each edge of a specific brain region. This representation highlights which regions mainly impact the classification.

### 3.3 Time scale of classification performance

We then focused on classification performance and the amount of signal used to compute the ATMs. As expected, the average classification performance increases as a function of the length of the segments. However, unlike the average, we note that the variance (across splits) of the accuracy does not become monotonically smaller as a function of the lengths of the segments. Instead, as one can observe in Fig.4, the variance of the classification is nearly flat for segments of 30s and of 180s. However, when one moves from 30 to 60 seconds, one can observe that the variance increases, since higher accuracies, that were never achieved with segments, can be reached in some splits. In other words, 30s saturates the classification performance, and to obtain improved classification one needs longer data segments.

**FIGURE 4.**
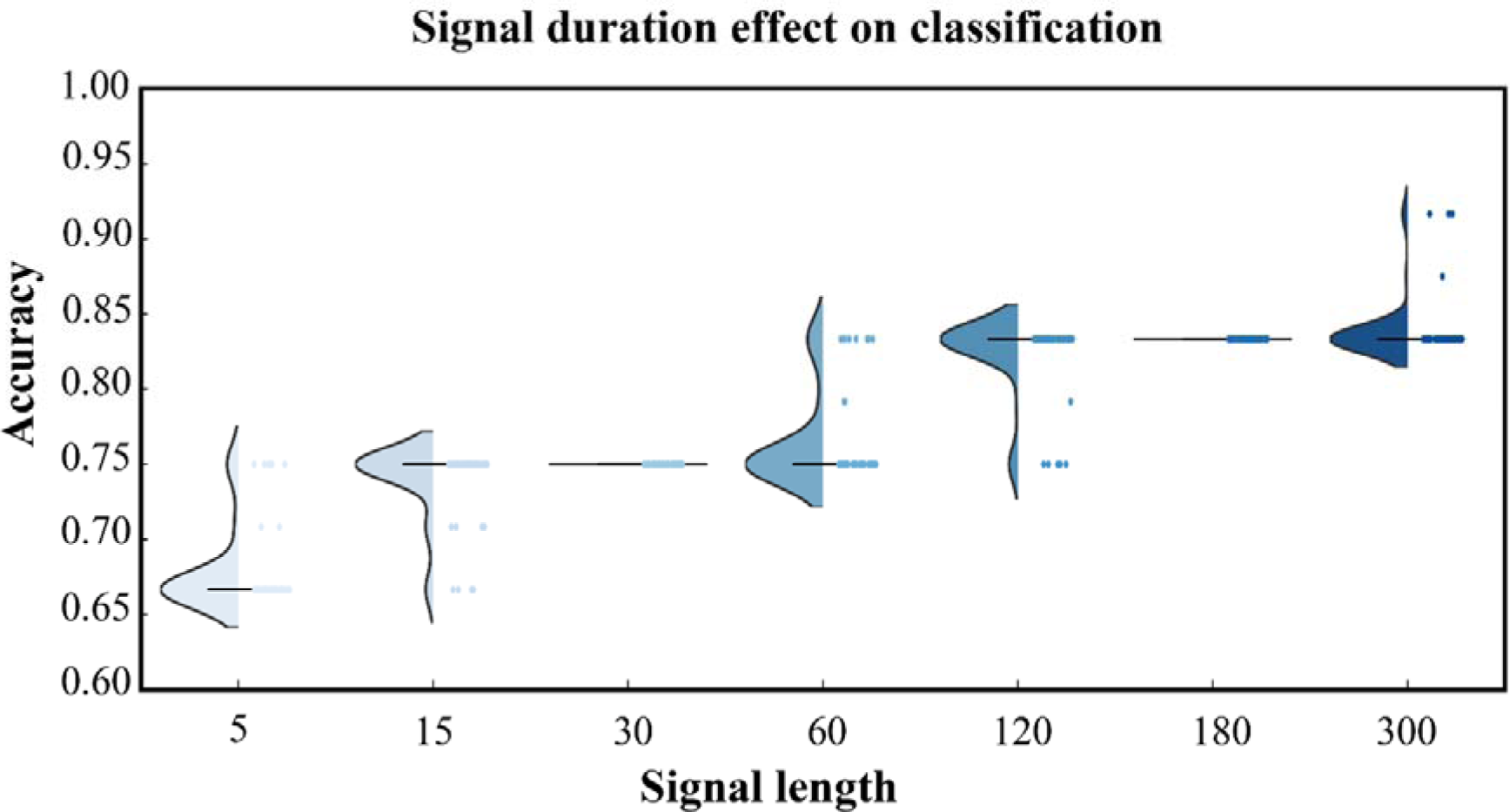
Time dependence of classification accuracy. The picture shows the distribution across splits of the cross-validation based on the amount of signal used to compute avalanche transition matrix (ATM). Each dot corresponds to the estimation of the classification performance per split.

## 4. Discussion

The present work aimed at improving the automatable classification of epileptic patients, utilizing machine learning on information of the brain dynamics derived no-invasively from hdEEG. To this purpose, we used the avalanche transition matrix (ATM) in order to measure how large-scale aperiodic perturbations spread across the brain. In particular, the ATMs are a recently developed analytical tool that captures the probability of any two regions being successively recruited by spontaneous neuronal avalanches, resulting in a functional network (Rucco et al., 2020; Sorrentino et al., 2021, 2021). Recently, we provided evidence that the ATMs are useful in temporal lobe epilepsy (TLE), as they allow the identification, from resting state hdEEG, of the regions with functional alterations, which are the ones generally involved in seizure initiation and propagation (Duma et al., 2023). As a first finding, the model classified subjects as patients with epilepsy or controls, with a mean accuracy of 0.87 ± 0.10(SD), with an AUC of 0.94 ± 0.06 (SD) and sensitivity of 0.89 ± 0.15 (SD) when using the ATMs-derived features. We observed that the model trained with ATMs outperforms the ImCoh across all the performance measures increasing the performance from 10 to 20% based on the measure of interest. The implications of this finding are twofold. From a theoretical perspective, the information conveyed by the large wave of aperiodic bursts is more informative and representative of the altered brain dynamics in TLE, as compared to the power-covariance-based connectivity measure (ImCoh). This can also be appreciated when observing the type of information that the model is using to perform classification. In the ATM-model, subsets of specific connections between brain nodes mainly drive the classification. Importantly, these edges are clustered into regions of fundamental relevance in TLE, which are typically involved both in seizure initiation and/or propagation, or characterized by well-known metabolic or connectivity alterations, such as the entorhinal cortex, the superior and inferior temporal gyri, the cingulate cortex, and the prefrontal dorsolateral cortex (Bernasconi et al., 1999; Jo et al., 2019; Kubota et al., 2013; Qin et al., 2020; Vismer et al., 2015). These findings suggest that the ATMs are a sensible measure, capable of capturing altered neural dynamics and characterizing the TLE patient. The better performance obtained with ATM as compared to the one with ImCoh, both in the broad- and narrow-band filtered signal, might suggest that the aperiodic components might be of relevance when focusing on the large-scale dynamics. In other words, the alterations are not limited to a single temporal scale, but rather affect multiple timescales. This is in line with emerging literature showing that multiple nested characteristic timescales characterize the brain dynamics at rest (Sorrentino, Rabuffo, et al., 2023). Along this line of evidence, the alterations induced by TLE might also affect multiple timescales. In fact, the classification accuracy saturates at specific durations of the time series, while it shows larger variations (across the particular segment taken into account for a patient) for intermediate lengths. In other words, for a length of 30 s, which particular segment has been used to perform the classification does not have any relevance, as the classification will have converged to a specific value. When one moves to segments of 60s, the situation is different, since the particular segment that is used for classification has an effect (the performance varies from one particular split to the next), which results in the variance of the classification performance across the segments. However, the average performance tends to rise as a function of the duration of the data segments, and never goes below the classification performance of the previous convergence time-point (i.e., 30s). At 120 seconds, the variance is similar, but the results of the classification start converging towards a new upper value, which is reached at 180 seconds. Indeed, at this duration, the classification performance converges again, and it will reach a specific value regardless of the specific segment. We propose that this might be an effect of the specific timescales that are affected by TLE. In other words, 30sec-long recordings would be enough to fully sample the faster dynamics. However, the latter are nested within slower dynamics which are affected by the TLE (Xie et al., 2023). For this reason, in order to exhaustively sample the brain dynamics complexity and exploit it for an efficient classification, longer segments are required. Furthermore, these results have a potential direct application in clinical practice. In fact, the rate of epilepsy misdiagnosis varies between 20-40% (Oto, 2017) and remains an issue also in specialized centers (Uldall et al., 2006). Some populations carry a particularly high risk of misdiagnosis, for example, patients with psychogenic non-epileptic seizures (Smith et al., 1999; Xu et al., 2016). Misidentification of EEG abnormalities reaches over 50% in the PNES population (Reuber et al., 2002). The method proposed in the present work may represent a potential tool for helping clinicians with diagnostics, which translates into the possibility of reducing the misinterpretation of EEG abnormalities by taking into account global brain activities. In fact, the present pipeline can be fully automated, requiring therefore minimal effort to the clinicians and the technical staff, that is the recording of a minimum of 5 minutes of resting state EEG. Importantly, the data have been analyzed without including epileptiform activities, which increases the generalizability of this analytical framework. In fact, in clinical practice, the identification of seizures and IEDs is a time-consuming process often performed manually. Moreover, seizures and/or IEDs are not always recorded, even during multiple days of monitoring. In this scenario, the possibility of classifying the patients using the intrinsic brain organization, without the necessity of epileptiform graphic elements, represents a strength of this approach. The present work does not aim to provide a tool to substitute the fundamental knowledge and experience of a clinical expert. On the contrary, this workflow should be considered as a potential user-friendly pipeline in support of clinicians in the diagnosis process, leveraging the time resolution and non-invasiveness of the EEG. The present pipeline might be of important help also in the low-income countries where patients have difficulty accessing specialist care (Dorji et al., 2023; Espinosa-Jovel et al., 2018). The manageability and cost-effectiveness of the EEG, together with the present workflow, may represent a suitable instrument for helping clinicians in their daily practice. In order to translate this pipeline to clinical practice, validation on multiple epilepsy and neurological populations is warranted. To summarize, our results provide evidence that the ATMs better capture the subtle neural dynamics alteration in epilepsy, as compared to classic functional connectivity measures, and that this may be effectively fed to artificial intelligence algorithms to improve automated classification of TLE patients.

## Data availability statement

The data that support the findings of this study are available on request to the corresponding author. The raw data are not publicly available due to privacy or ethical restrictions. All the scripts are available at the following github page: https://github.com/mccorsi/NeuronalAvalanches_TemporalLobeEpilepsy_EEG.

## Supplementary materials

**Supplementary Figure 1.**
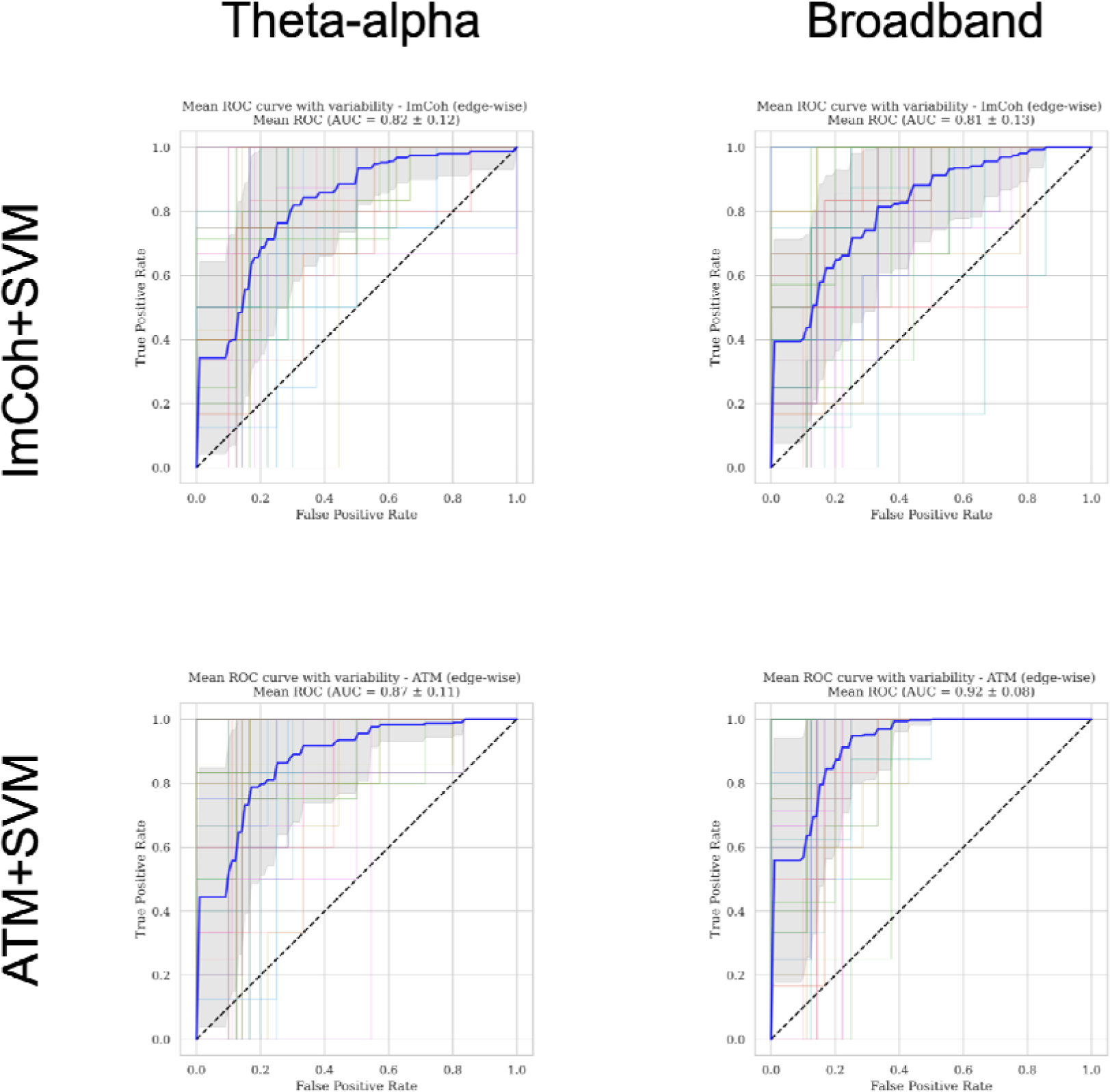
ROC Curves obtained with ImCoh+SVM and ATM+SVM within the broadband and the narrowband (theta-alpha) signal. Each figure represents the distribution of the ROC Curves across the splits and the blue curve corresponds to the mean of the curves.

**Supplementary Figure 2.**
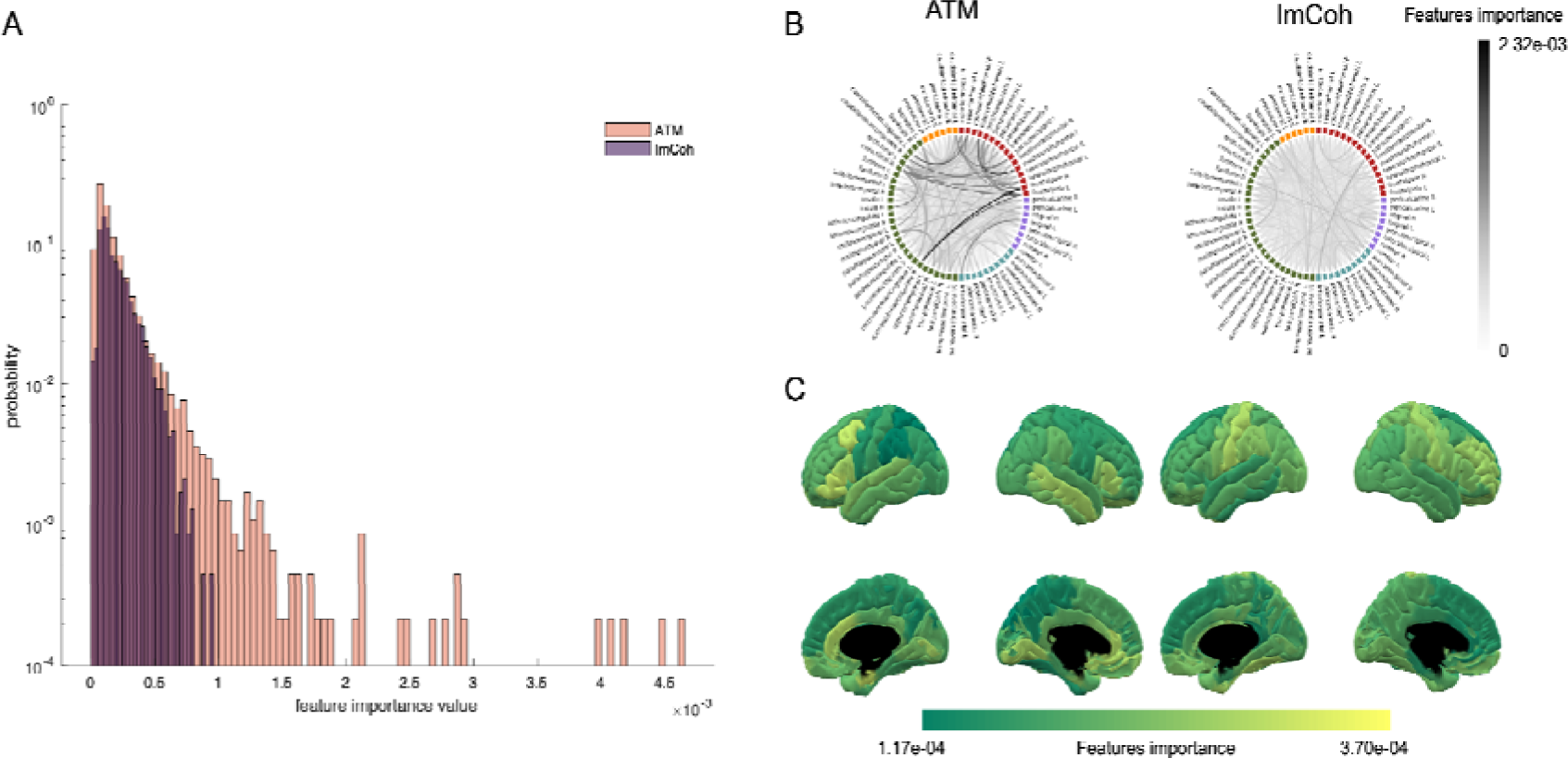
Feature importance for model interpretability (narrow band). The present figure represents the relevance of the information used by the model to perform classification, namely the feature importance, both using avalanche transition matrix (ATM) and imaginary coherence (ImCoh). Panel A is a histogram of the probability of the feature importance value in ATM and ImCoh. The histogram shows a narrow distribution for ImCoh and broader distribution for ATM, suggesting that in ATM that certain edges drive the majority of information necessary for differentiation of the two groups. Panel B is the edge representation in a chord plot, showing the importance of each edge in the classification. Finally, panel C is the mean importance value of each edge of a specific brain region. This representation highlights which regions mainly impact in the classification.

**Supplementary Figure 3.**
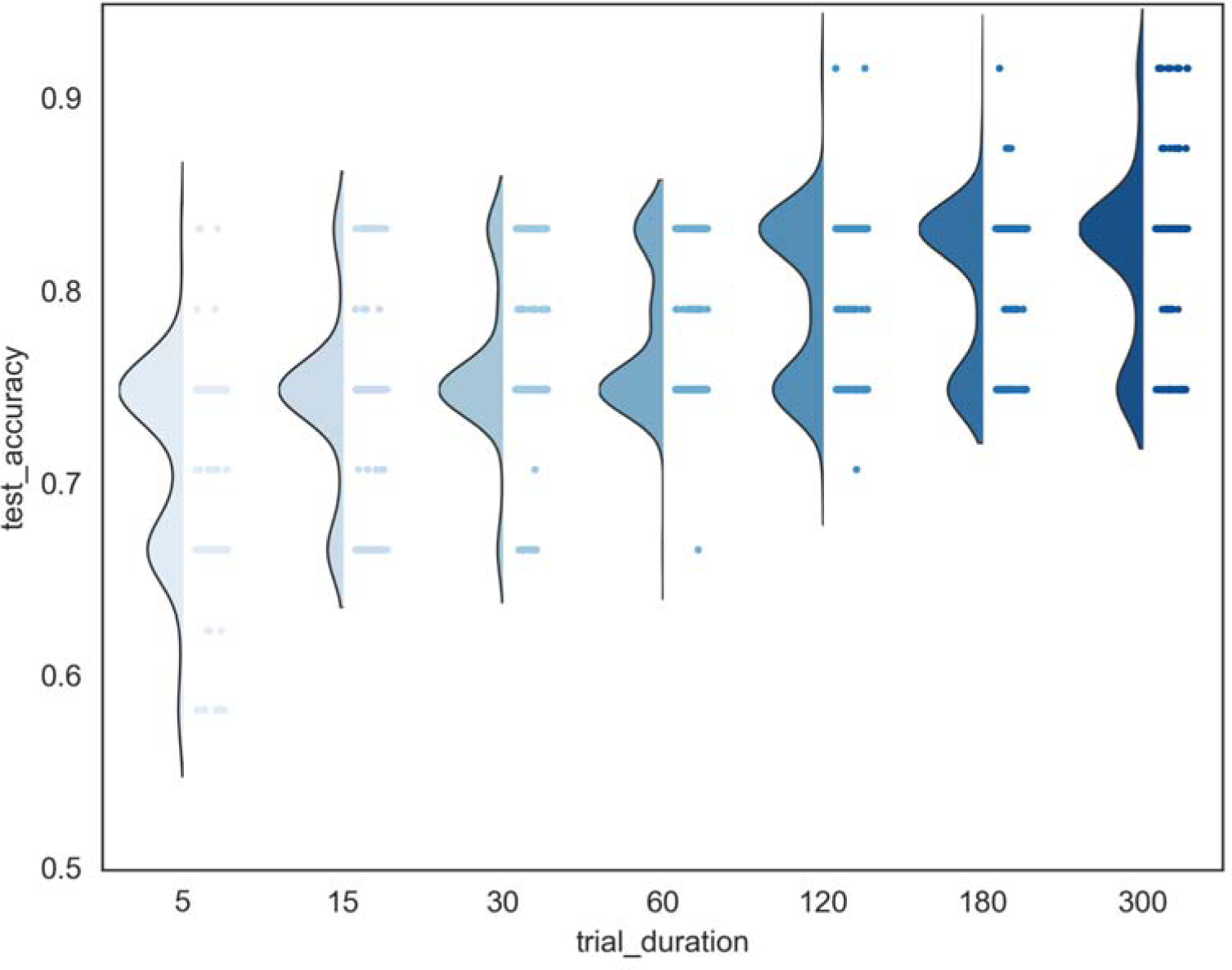
Time dependence of classification accuracy - narrow band. The picture shows the distribution across splits of the cross-validation based on the amount of signal used to compute avalanche transition matrix (ATM).

